# Examining the Etiological Specificity of Genetic Risk for Alcohol Use Disorder and Alcohol Consumption through Symptom-Level Mediation Analyses

**DOI:** 10.64898/2026.10.28.26366761

**Authors:** Yongguk M. Sarles, Alex P. Miller, Joseph D. Deak, Kirk C. Wilhelmsen, Ian R. Gizer

**Affiliations:** Department of Psychological Sciences, University of Missouri-Columbia; Department of Psychiatry, Indiana University School of Medicine; Department of Psychiatry, Yale University School of Medicine; Veterans Affairs Connecticut Healthcare System; Rockefeller Neuroscience Institute, West Virginia University; Department of Neurology, West Virginia University

**Keywords:** alcohol use disorder, polygenic score, mediation analysis, personality psychopathology, alcohol consumption, genetic risk

## Abstract

**Introduction:** Polygenic scores (PGSs) for alcohol use disorder (AUD) may capture genetic liability related to alcohol consumption as well as broader transdiagnostic traits, including impulsivity, aggression, and neuroticism. However, the extent to which these traits account for associations between an AUD PGS and the presentation of AUD symptoms remains unclear. Thus, to better understand the genetic influences captured by an AUD PGS, the present study used mediation analyses to examine whether these transdiagnostic traits differentially mediate the relations between PGSs for AUD and individual AUD symptoms while conducting a parallel analysis using a PGS of alcohol consumption to examine differences in the genetic influences underlying AUD and consumption.

**Method:** Participants were drawn from the UCSF Family Alcoholism Study. Separate mediation models examined maximum habitual drinks (*N* = 1,611) and MMPI-2 Personality Psychopathology Five (PSY-5) scales (*N* = 1,464) as mediators of the association between AUD/alcohol consumption PGSs and individual AUD symptoms. Covariates included sex, age, and 10 ancestry principal components. Observations were modeled as clustered within families, and a Bonferroni correction was applied (*p* < .0045).

**Results:** Relations between the AUD PGS and AUD symptoms were fully mediated by maximum habitual drinks and partially mediated by neuroticism for nearly all symptoms, but not by other PSY-5 scales. Relations between the alcohol consumption PGS and AUD symptoms were fully mediated by maximum habitual drinks alone, with no mediation observed through any personality psychopathology measure.

**Discussion:** Current AUD and alcohol consumption PGSs appear to capture genetic liability that operates predominantly through alcohol consumption. However, the AUD PGS may capture genetic influences on broader transdiagnostic factors as well, with neuroticism acting as a partial mediator. Thus, etiological interpretations of the AUD PGS as solely a measure of AUD risk should be made cautiously.

## Introduction

Alcohol use disorder (AUD) is a heritable psychiatric condition, with twin and adoption studies converging on a heritability estimate of roughly 50% (Verhulst et al., 2015). Genome-wide association studies (GWAS) have since begun to identify the specific genetic variants behind this heritability, and polygenic scores (PGSs) built from these GWAS are now widely used as quantitative indices of an individual’s genetic liability to AUD. This work has made clear that genetic liability to AUD is not simply a liability to drink heavily (Mallard et al., 2022; Walters et al., 2018). Rather, it appears to reflect an interplay of alcohol metabolism and consumption as well as broader, transdiagnostic risk factors, consistent with current models of AUD etiology (e.g., Bogdan et al., 2023; Boness et al., 2021).

At a minimum, an AUD PGS is expected to capture genetic influences on alcohol consumption. Alcohol use itself is not part of the DSM-5 criteria, but it is a necessary precondition for the disorder: a person cannot develop AUD without drinking, which makes it difficult for genetic studies to fully separate diagnosis-specific genetic liability from genetic liability to consumption itself. Several lines of genomic evidence point in this direction. Genetic correlations between AUD diagnosis and alcohol consumption are moderate, indicating that the two are genetically related, but not identical (Kranzler et al., 2019; Walters et al., 2018). Further, AUD shows substantial positive genetic correlations with multiple forms of psychopathology and reduced social-emotional functioning whereas alcohol consumption shows reduced or even negative correlations with these same phenotypes (Kember et al., 2023; Kranzler et al., 2019). In addition, multivariate genomic analyses indicate that genetic liability to problematic alcohol use can be decomposed into a component shared with broader externalizing liability and a component specific to alcohol use (Barr et al., 2022). Despite such findings, few studies have examined how the different sources of genetic liability reflected in these genetic correlations might be reflected in a PGS for AUD (e.g., Ksinan et al., 2019; Li et al., 2017; Su et al., 2021).

Contemporary models of AUD etiology identify specific constructs relevant to cognition and personality as central facets of AUD risk (Bogdan et al., 2023; Boness et al., 2021). For example, behavioral disinhibition has been consistently described as one such risk factor that provides a pathway to the disorder (Sher et al., 2005). This behavioral disinhibition or impulsivity pathway has been formalized in the externalizing spectrum model, which conceptualizes substance use disorders, antisocial behavior, and impulsive and aggressive personality traits as indicators of a single, highly heritable latent liability (Karlsson Linnér et al., 2021; Krueger et al., 2009). Neuroticism represents a second, partially distinct personality dimension long implicated in AUD risk, typically through pathways involving negative affect regulation and drinking to cope (Kuntsche et al., 2006; Littlefield et al., 2011). Twin-based behavior genetic studies have consistently shown that alcohol problem measures share genetic variance with neuroticism (De Moor et al., 2011; Littlefield et al., 2011; Slutske et al., 2002), and large-scale GWAS of alcohol dependence have confirmed that its genetic architecture correlates with a range of psychiatric and behavioral traits, including neuroticism (Zhou et al., 2023). Given this literature, we expect that AUD genetic liability, if it in part indexes transdiagnostic liability as contemporary models of AUD etiology suggest, should act partially through liability toward impulsivity and aggression as well as neuroticism.

Though few, molecular genetics studies of these pathways have begun using PGS methods to examine how genetic risk for alcohol use and alcohol problems may be expressed. For example, Li et al. (2017) found that the association between a PGS for alcohol use problems and adolescent alcohol involvement was partially explained by sensation seeking, indicating a mediating role for impulsivity in how genetic liability is expressed. Subsequent studies have reported similar mediating roles for sensation seeking along with related impulsivity facets (e.g., lack of perseverance) in college and adult samples (Ksinan et al., 2019; Su et al., 2021). Despite the described genetic correlations, to our knowledge, only candidate gene studies have been conducted evaluating the role of neuroticism in the pathway from genetic risk to alcohol problems (Cai et al., 2024; Criado et al., 2014). Thus, further studies are needed to understand the pathways through which this genetic risk is expressed.

One potentially fruitful method for investigating these pathways is to explore whether they contribute similarly to each of the AUD diagnostic symptoms. Symptom-level analyses are critical given that AUD, as defined by the DSM-5, is a polythetic diagnosis comprising 11 criteria that span physiological (e.g., tolerance, withdrawal), behavioral (e.g., hazardous use, activities given up), and social (e.g., interpersonal problems) domains (American Psychiatric Association, 2013). More recent studies have characterized the suggested heterogeneity in etiologies that may exist across such symptom clusters, including at the genetic level (Kendler et al., 2012; Watts et al., 2021). In addition, researchers have identified shared and symptom-specific additive genetic effects on DSM-5 criteria (Palmer et al., 2019) and distinct genome-wide significant loci for individual criteria and criterion counts (Lai et al., 2019), a pattern also observed in item-level GWAS of related AUD screening measures (Mallard et al., 2022). Lastly, Sarles and colleagues (2025) showed that this heterogeneity is also evident in the differential genetic severity of individual AUD symptoms. Diagnosis or symptom-count phenotypes, of the kind typically used as GWAS discovery phenotypes, necessarily average over this heterogeneity and may obscure whether genetic risk operates uniformly across symptoms or is concentrated in a subset of criteria more closely tied to consumption. Modeling each symptom separately therefore may offer a more nuanced test of etiological specificity than modeling AUD diagnosis or symptom count as a single outcome.

The reviewed studies suggest that symptom-level investigations of AUD genetic risk are warranted. Examining a PGS’s relation to individual AUD symptoms offers an opportunity to identify which underlying processes account for the genetic influences captured by AUD GWAS, and whether those processes differ from one symptom to the next. We therefore tested alcohol consumption and personality psychopathology, indexed by the MMPI-2 Personality Psychopathology Five scales (PSY-5; Harkness et al., 1995, 2012), as candidate mediators of the relation between an AUD PGS and each of the 11 AUD symptoms. Specifically, we expected that the AUD PGS’s relation to each symptom would be explained not only by alcohol consumption but by personality psychopathology as well, such as impulsivity, aggression, and neuroticism.

Because it was unclear at the outset whether these mediation patterns would generalize beyond genetic liability captured by a diagnosis-based phenotype, we planned, in parallel, an analogous set of mediation models using a PGS for drinks per week (DPW), derived from a substantially larger, consumption-specific GWAS (Saunders et al., 2022), applied to the same sample, the same 11 AUD symptoms, and the same set of candidate mediators. This design choice was motivated by two considerations. First, if the AUD PGS showed the expected transdiagnostic pattern of mediation, the DPW PGS would provide an informative specificity check, given that a purely consumption-based phenotype should, in principle, show a narrower pattern of mediation restricted to alcohol consumption itself. Second, molecular genetic studies have reported nontrivial genetic correlations between alcohol consumption and impulsive personality traits assessed via the UPPS-P (Sanchez-Roige et al., 2023; Stephenson et al., 2023).

Given this documented genome-wide genetic correlation between consumption phenotypes and impulsivity, it was plausible that the DPW PGS, despite being nominally a narrower, consumption-specific score, would nonetheless show mediation through impulsivity at the phenotypic level. We therefore tested this possibility directly alongside the analogous models for the AUD PGS.

## Method

### Discovery samples

The AUD PGS was calculated from GWAS summary statistics derived from a sample-size-weighted meta-analysis of three discovery cohorts, originally reported by Miller & Gizer (2024). These cohorts included a Psychiatric Genomics Consortium GWAS of DSM-IV alcohol dependence with an effective *N* of 26,853 (Walters et al., 2018), a FinnGen Consortium Release 6 GWAS of AUD with an effective *N* of 40,997 (Kurki et al., 2022), and a Million Veteran Program GWAS of AUD with an effective *N* of 152,332 (Zhou et al., 2020). The three sets of summary statistics were combined using a sample-size-weighted meta-analysis in METAL (Willer et al., 2010) for variants with a minor allele frequency greater than 0.01 (9,533,157 variants), restricted to participants of European-like ancestry, yielding a combined effective *N* of 220,182.

A parallel PGS was created for DPW to evaluate whether a purer consumption phenotype would show a different pattern of mediation. The DPW PGS was calculated from summary statistics from a meta-analysis of self-reported drinks consumed per week, with the 23andMe dataset removed, with an effective *N* of 2,965,643 (Saunders et al., 2022).

### Target sample

Participants were drawn from the University of California, San Francisco (UCSF) Family Alcoholism Study, a community-ascertained family study designed to identify genetic loci that influence susceptibility to alcohol dependence and related phenotypes (Vieten et al., 2004). The UCSF Family Study recruited proband participants who met lifetime alcohol dependence criteria (*ICD-10*, 2004) and their available family members. Analyses were restricted to participants of European-like ancestry because only a small number of non-European-like ancestry participants were enrolled in the UCSF study (approximately 96% of participants were of European-like ancestry). Furthermore, the predictive accuracy of PGSs is reduced when the ancestral background of the target sample differs from that of the discovery GWAS (Martin et al., 2017; Otto et al., 2016). Participants who had never been exposed to alcohol were also excluded.

Demographics and descriptive statistics for all study variables, including means, standard deviations, and ranges, are presented in Table 1. Institutional review board committees approved all data collection, and participants provided informed consent prior to participation.

**Table 1.**
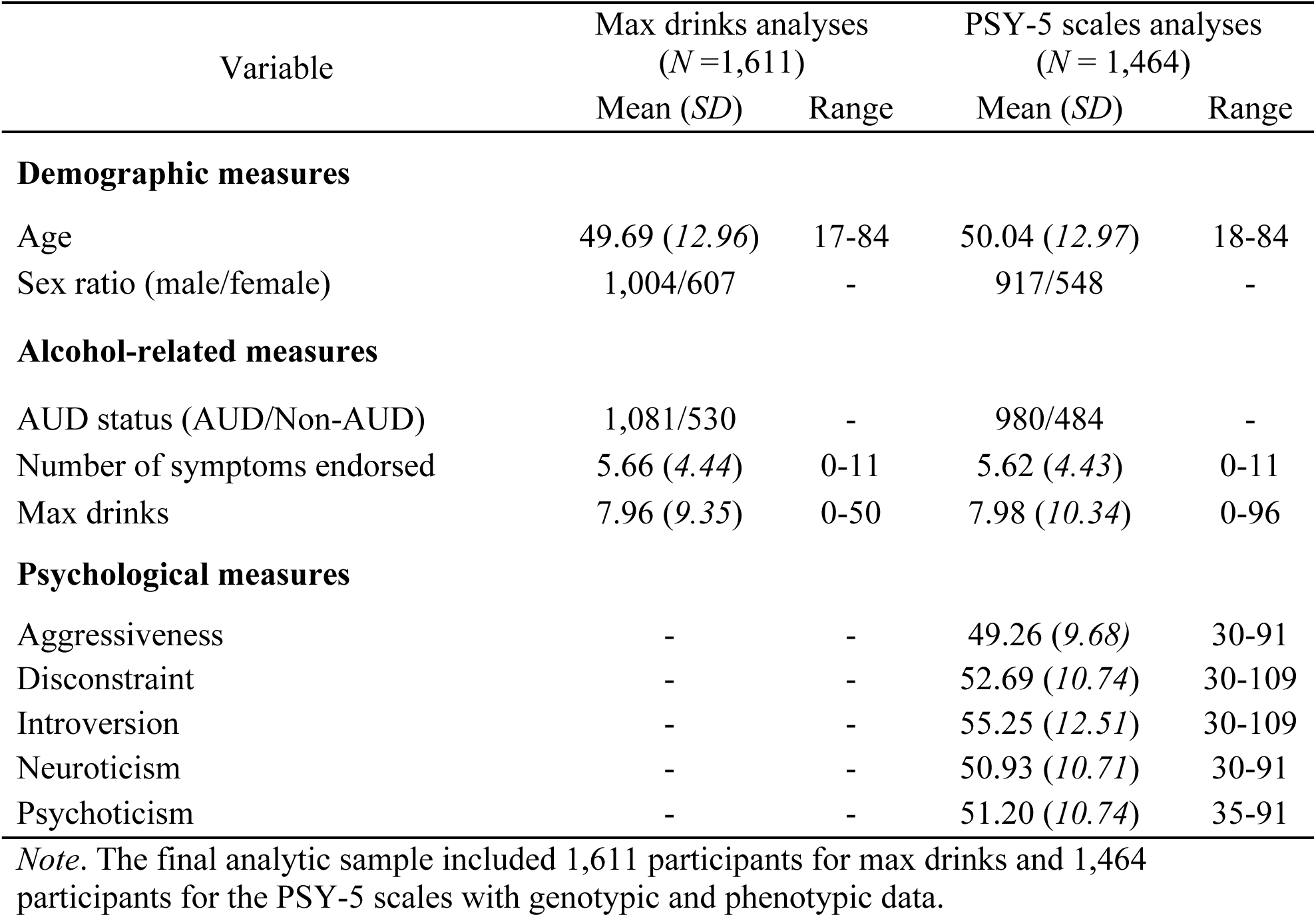
Demographic Characteristics and Descriptive Statistics for Study Variables.

#### Measures

##### AUD symptoms

The 11 DSM-5 AUD symptom criteria were approximated using items from a modified version of the Semi-Structured Assessment for the Genetics of Alcoholism (SSAGA), a comprehensive psychiatric interview developed for use in genetically informed studies of alcoholism (Bucholz et al., 1994). For example, craving was assessed with the item, “In situations where you couldn’t drink, did you ever have such a strong desire for it that you couldn’t think of anything else.” Symptoms included *tolerance, craving, larger/longer, cut/stop, role failure, interpersonal problems, time spent, reduced activities, withdrawal, hazardous use, and continued use (despite physical/psychological problems)*.

##### Alcohol consumption

Alcohol consumption was operationalized as the maximum habitual alcohol intake in 24 hours during the period of the participant’s heaviest drinking (max drinks), also assessed by the SSAGA (i.e., “What was the largest number of drinks you would drink almost every day?”). Participants with outlying alcohol consumption values (i.e., maximum habitual drinks [max drinks]; > 60) were excluded from analyses.

##### Personality psychopathology

Personality pathology was assessed using the MMPI-2 PSY-5 scales, a dimensional descriptive system for personality and its disorders comprising the domains of aggressiveness, disconstraint (i.e., impulsivity), introversion/low positive emotionality, neuroticism/negative emotionality, and psychoticism (Harkness et al., 1995, 2012).

#### Whole genome sequencing

Sequencing and genotyping methods for the UCSF Family Study have been described previously (Gizer et al., 2018). Sequencing was performed using HiSeq2000 sequencers (Illumina, San Diego, CA), with the majority of participants sequenced at a coverage depth of 2x to 6x. Sequence variants were called using Thunder, an LD-aware variant-calling method (Li et al., 2011). Participants were also genotyped using Affymetrix microarrays to assess the quality of sequence-derived genotype calls, with comparisons between the two platforms showing 98% concordance. Ancestry principal component analysis (PCA; Price et al., 2010) was conducted in Genome-wide Complex Trait Analysis (GCTA; Yang et al., 2011) using sequenced variants with a minor allele frequency greater than 0.01.

#### Polygenic scores (PGS)

PGSs for the AUD and DPW discovery phenotypes were generated using PRS-CS, a Bayesian regression method that places continuous shrinkage priors on single nucleotide polymorphism effect sizes and models local linkage disequilibrium patterns using an external reference panel, without requiring hard clumping and thresholding of the discovery summary statistics (Ge et al., 2019). European-like ancestry samples from the 1000 Genomes Project (phase 3, NCBI GRCh37) served as the linkage disequilibrium reference panel, and only variants present across the discovery, reference, and UCSF datasets were retained. Final PGSs were constructed by summing and standardizing weighted allele counts (mean = 0, *SD* = 1) using PLINK 2.0 (Chang et al., 2015).

### Data analysis

For each PGS, separate single-mediator models were estimated for each of the 11 AUD symptoms to evaluate the indirect effect of the PGS acting through max drinks and each personality psychopathology scale. Because each symptom was a binary outcome, models were estimated with the weighted least squares means and variance adjusted (WLSMV) estimator.

Indirect effects were estimated as the product of the path from the PGS to the mediator and the path from the mediator to the AUD symptom, and their statistical significance was tested with delta-method standard errors. Mediation was considered full when the direct effect of the PGS on the symptom was reduced to non-significance after accounting for the mediator, and partial when the direct effect remained significant alongside a significant indirect effect. Sex, age, and 10 ancestry principal components were included as covariates in each model, and observations were modeled as clustered within families to account for the nonindependence of relatives (see Figure 1 for an illustration). A Bonferroni correction for multiple testing was applied across the full set of mediation tests to control the family-wise error rate (0.05/11 = 0.0045; *p* < .0045). All analyses were performed using Mplus version 8 (Muthén & Muthén, 2017). The analyses were not preregistered. Therefore, the findings should be interpreted as exploratory.

**Figure 1.**
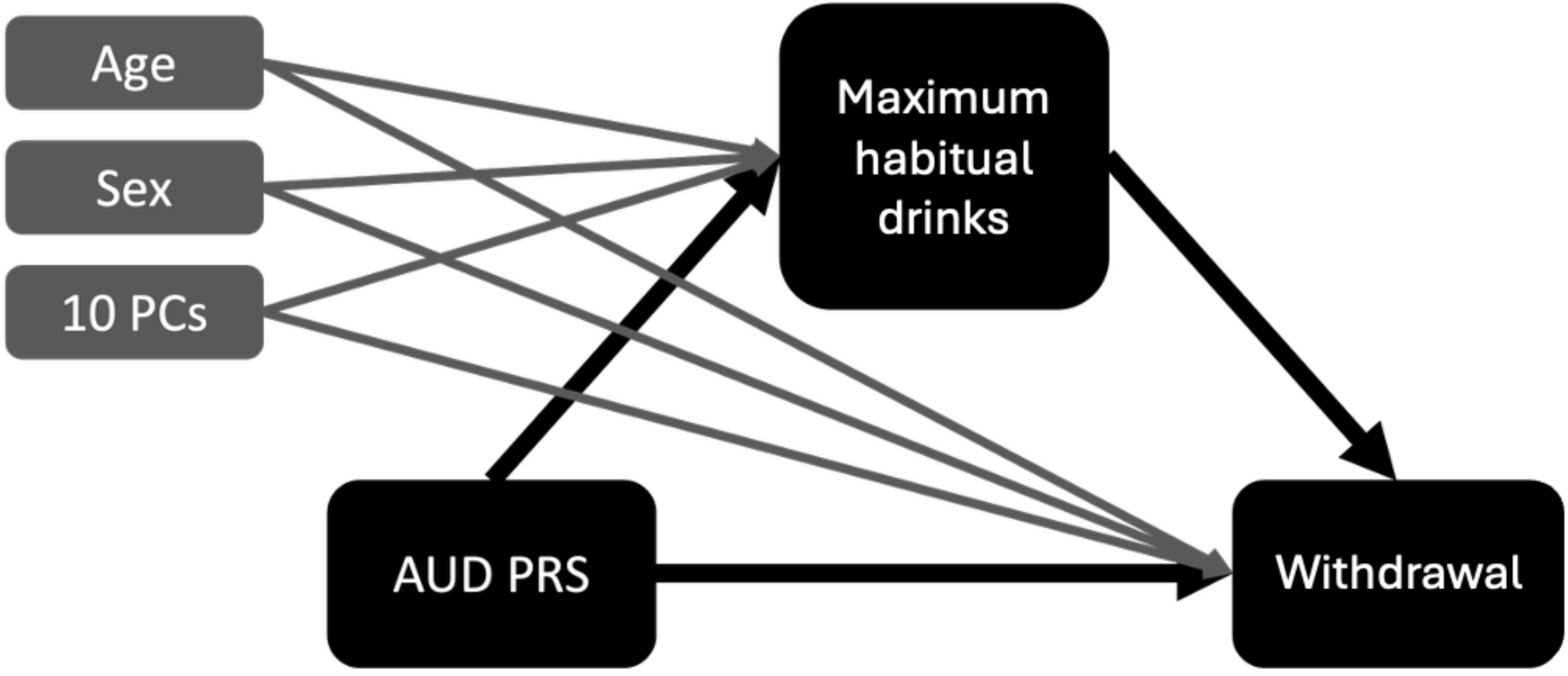
Example of Mediation Analyses. *Note.* For simplicity, only the path from AUD PGS to *withdrawal* through maximum habitual drinks was included in this example. Sex, age, and 10 ancestry principal components were included as covariates in each model. Observations were modeled as clustered within families to address the nonindependence among observations.

## Results

### AUD PGS

Relations between the AUD PGS and individual AUD symptoms were fully mediated by max drinks for 10 of the 11 symptoms (indirect effect *ps* < .001, direct effect *ps* > .009). The exception was *withdrawal*, for which mediation was only partial (indirect effect *p* < .001, direct effect *p* = .003). Neuroticism acted as a partial mediator for most symptoms (indirect effect *ps* < .003, direct effect *ps* < .003). The exceptions were *Cut/Stop* and *Hazardous Use*, for which neuroticism fully mediated the relation with the AUD PGS (indirect effect *p* = .002, direct effect *p* = .012).

As exploratory analyses, we conducted parallel mediation analyses where neuroticism and max drinks were included simultaneously as mediators. Max drinks remained a significant mediator of the relations between the AUD PGS and all AUD symptoms, consistent with the single-mediator results. Neuroticism’s indirect effects, by contrast, were substantially attenuated once max drinks was included in the same model, remaining significant only for *craving, tolerance, and reduced activities*. Full single-mediator results of the AUD PGS are presented in Table 2. The unique associations between each mediator and individual AUD symptoms after accounting for the AUD PGS as well as results of the exploratory parallel mediation analyses, are presented in Supplementary Tables 1 and 2.

**Table 2.** Results of Mediation Analyses for the AUD Polygenic Score.

| Dependent variable | Mediators |  |  |  |  |  |  |  |  |  |  |  |
| --- | --- | --- | --- | --- | --- | --- | --- | --- | --- | --- | --- | --- |
|  | Maximum habitual drinks |  | Aggressiveness |  | Disconstraint |  | Neuroticism |  | Introversion |  | Psychoticism |  |
|  | Indirect effect<br>(*p*) | Direct effect<br>(*p*) | Indirect effect<br>(*p*) | Direct effect<br>(*p*) | Indirect effect<br>(*p*) | Direct effect<br>(*p*) | Indirect effect<br>(*p*) | Direct effect<br>(*p*) | Indirect effect<br>(*p*) | Direct effect<br>(*p*) | Indirect effect<br>(*p*) | Direct effect<br>(*p*) |
| Cut/Stop | **0.071\***<br>(*< .001*) | 0.033<br>(*0.280*) | 0.004<br>(*0.485*) | **0.101\***<br>(*0.002*) | 0.008<br>(*0.336*) | **0.098\***<br>(*0.001*) | **0.024\***<br>(*0.002*) | 0.082<br>(*0.012*) | 0.001<br>(*0.638*) | **0.105\***<br>(*0.001*) | 0.005<br>(*0.270*) | **0.101\***<br>(*0.002*) |
| Larger /longer | **0.076\***<br>(*< .001*) | 0.054<br>(*0.085*) | 0.005<br>(*0.486*) | **0.127\***<br>(*< .001*) | 0.011<br>(*0.336*) | **0.121\***<br>(*< .001*) | **0.027\***<br>(*0.001*) | **0.105\***<br>(*0.002*) | 0.001<br>(*0.641*) | **0.131\***<br>(*< .001*) | 0.005<br>(*0.270*) | **0.127\***<br>(*< .001*) |
| Craving | **0.061\***<br>(*< .001*) | 0.076<br>(*0.010*) | 0.003<br>(*0.486*) | **0.144\***<br>(*< .001*) | 0.007<br>(*0.337*) | **0.140\***<br>(*< .001*) | **0.026\***<br>(*0.001*) | **0.121\***<br>(*< .001*) | 0.001<br>(*0.642*) | **0.146\***<br>(*< .001*) | 0.005<br>(*0.266*) | **0.142\***<br>(*< .001*) |
| Time spent | **0.066\***<br>(*< .001*) | 0.052<br>(*0.073*) | 0.005<br>(*0.486*) | **0.121\***<br>(*< .001*) | 0.009<br>(*0.335*) | **0.118\***<br>(*< .001*) | **0.025\***<br>(*0.001*) | **0.101\***<br>(*< .001*) | 0.001<br>(*0.635*) | **0.125\***<br>(*< .001*) | 0.005<br>(*0.265*) | **0.121\***<br>(*< .001*) |
| Tolerance | **0.081\***<br>(*< .001*) | 0.054<br>(*0.086*) | 0.006<br>(*0.485*) | **0.134\***<br>(*< .001*) | 0.012<br>(*0.334*) | **0.129\***<br>(*< .001*) | **0.031\***<br>(*0.001*) | **0.109\***<br>(*0.001*) | 0.001<br>(*0.634*) | **0.139\***<br>(*< .001*) | 0.006<br>(*0.270*) | **0.134\***<br>(*< .001*) |
| Activities given up | **0.060\***<br>(*< .001*) | 0.073<br>(*0.015*) | 0.003<br>(*0.488*) | **0.128\***<br>(*< .001*) | 0.008<br>(*0.336*) | **0.124\***<br>(*< .001*) | **0.027\***<br>(*0.001*) | **0.105\***<br>(*0.001*) | 0.002<br>(*0.633*) | **0.130\***<br>(*< .001*) | 0.005<br>(*0.267*) | **0.126\***<br>(*< .001*) |
| Role failure | **0.060\***<br>(*< .001*) | 0.064<br>(*0.030*) | 0.004<br>(*0.487*) | **0.126\***<br>(*< .001*) | 0.008<br>(*0.335*) | **0.121\***<br>(*< .001*) | **0.026\***<br>(*0.001*) | **0.103\***<br>(*0.001*) | 0.002<br>(*0.636*) | **0.128\***<br>(*< .001*) | 0.005<br>(*0.266*) | **0.125\***<br>(*< .001*) |
| Withdrawal | **0.068\***<br>(*< .001*) | **0.085\***<br>(*0.003*) | 0.004<br>(*0.489*) | **0.158\***<br>(*< .001*) | 0.009<br>(*0.336*) | **0.153\***<br>(*< .001*) | **0.026\***<br>(*0.002*) | **0.136\***<br>(*< .001*) | 0.001<br>(*0.636*) | **0.160\***<br>(*< .001*) | 0.006<br>(*0.267*) | **0.156\***<br>(*< .001*) |
| Continued use | **0.081\***<br>(*< .001*) | 0.058<br>(*0.072*) | 0.006<br>(*0.484*) | **0.135\***<br>(*< .001*) | 0.014<br>(*0.334*) | **0.127\***<br>(*< .001*) | **0.029\***<br>(*0.001*) | **0.112\***<br>(*0.001*) | 0.001<br>(*0.640*) | **0.140\***<br>(*< .001*) | 0.006<br>(*0.269*) | **0.135\***<br>(*< .001*) |
| Hazardous use | **0.071\***<br>(*< .001*) | 0.033<br>(*0.319*) | 0.005<br>(*0.485*) | **0.106\***<br>(*0.002*) | 0.012<br>(*0.334*) | **0.099\***<br>(*0.003*) | **0.025\***<br>(*0.002*) | 0.086<br>(*0.012*) | 0.001<br>(*0.640*) | **0.110\***<br>(*0.001*) | 0.005<br>(*0.271*) | **0.106\***<br>(*0.002*) |
| Interpersonal problems | **0.080\***<br>(*< .001*) | 0.057<br>(*0.074*) | 0.006<br>(*0.485*) | **0.133\***<br>(*< .001*) | 0.013<br>(*0.334*) | **0.126\***<br>(*< .001*) | **0.028\***<br>(*0.001*) | **0.111\***<br>(*0.001*) | 0.001<br>(*0.648*) | **0.138\***<br>(*< .001*) | 0.005<br>(*0.271*) | **0.134\***<br>(*< .001*) |

Note. Sex, age, and 10 ancestry principal components were included as covariates. Observations were modeled as clustered within families. A Bonferroni correction for multiple testing was applied ( $p < .0045$ ). \* $p < .0045$

Contrary to our hypothesis that impulsivity and aggression would mediate the relations between the AUD PGS and individual AUD symptoms, no significant mediation effects were observed for aggressiveness, disconstraint, introversion, or psychoticism (indirect effect *ps* > .265). These traits nonetheless showed unique associations with AUD symptoms that were independent of the AUD PGS, indicating that impulsivity and aggression may relate to AUD symptomatology through pathways that are not well captured by the genetic variance indexed by the current AUD PGS.

### DPW PGS

As with the AUD PGS, relations between the DPW PGS and individual AUD symptoms were fully mediated by max drinks, and this pattern held for all 11 symptoms, including *withdrawal* (indirect effect *ps* < .001, direct effect *ps* > .023). Full single-mediator results of the DPW PGS are shown in Table 3. The unique associations between each mediator and individual AUD symptoms after accounting for the DPW PGS are presented in Supplementary Table 3.

**Table 3.** Results of Mediation Analyses for the DPW Polygenic Score.

| Dependent variable | Mediators |  |  |  |  |  |  |  |  |  |  |  |
| --- | --- | --- | --- | --- | --- | --- | --- | --- | --- | --- | --- | --- |
|  | Maximum habitual drinks |  | Aggressiveness |  | Disconstraint |  | Neuroticism |  | Introversion |  | Psychoticism |  |
|  | Indirect effect<br>(*p*) | Direct effect<br>(*p*) | Indirect effect<br>(*p*) | Direct effect<br>(*p*) | Indirect effect<br>(*p*) | Direct effect<br>(*p*) | Indirect effect<br>(*p*) | Direct effect<br>(*p*) | Indirect effect<br>(*p*) | Direct effect<br>(*p*) | Indirect effect<br>(*p*) | Direct effect<br>(*p*) |
| Cut/Stop | **0.083\***<br>(*< .001*) | 0.024<br>(0.403) | 0.009<br>(0.126) | **0.098\***<br>(0.001) | 0.019<br>(0.014) | **0.088\***<br>(0.003) | 0.007<br>(0.316) | **0.100\***<br>(0.001) | 0.001<br>(0.769) | **0.107\***<br>(0.001) | 0.003<br>(0.270) | **0.104\***<br>(0.001) |
| Larger /longer | **0.090\***<br>(*< .001*) | 0.030<br>(0.328) | 0.011<br>(0.120) | **0.109\***<br>(0.001) | 0.029<br>(0.012) | **0.091\***<br>(0.004) | 0.008<br>(0.313) | **0.112\***<br>(*< .001*) | 0.001<br>(0.770) | **0.119\***<br>(*< .001*) | 0.003<br>(0.475) | **0.117\***<br>(*< .001*) |
| Craving | **0.072\***<br>(*< .001*) | 0.056<br>(0.045) | 0.007<br>(0.127) | **0.121\***<br>(*< .001*) | 0.016<br>(0.016) | **0.111\***<br>(*< .001*) | 0.007<br>(0.313) | **0.120\***<br>(*< .001*) | 0.000<br>(0.771) | **0.127\***<br>(*< .001*) | 0.003<br>(0.471) | **0.124\***<br>(*< .001*) |
| Time spent | **0.078\***<br>(*< .001*) | -0.003<br>(0.913) | 0.012<br>(0.111) | 0.063<br>(0.045) | 0.023<br>(0.013) | 0.052<br>(0.091) | 0.007<br>(0.316) | 0.068<br>(0.031) | 0.001<br>(0.768) | 0.074<br>(0.021) | 0.004<br>(0.469) | 0.071<br>(0.025) |
| Tolerance | **0.095\***<br>(*< .001*) | 0.015<br>(0.602) | 0.013<br>(0.115) | **0.097\***<br>(0.001) | 0.029<br>(0.012) | **0.081\***<br>(0.006) | 0.009<br>(0.313) | **0.101\***<br>(0.001) | 0.001<br>(0.768) | **0.109\***<br>(0.001) | 0.004<br>(0.471) | **0.106\***<br>(0.001) |
| Activities given up | **0.070\***<br>(*< .001*) | 0.017<br>(0.537) | 0.007<br>(0.121) | 0.080<br>(0.009) | 0.020<br>(0.014) | 0.067<br>(0.026) | 0.008<br>(0.314) | 0.080<br>(0.009) | 0.001<br>(0.767) | 0.086<br>(0.005) | 0.003<br>(0.468) | 0.084<br>(0.007) |
| Role failure | **0.070\***<br>(*< .001*) | 0.008<br>(0.782) | 0.007<br>(0.120) | 0.071<br>(0.024) | 0.021<br>(0.014) | 0.057<br>(0.060) | 0.007<br>(0.314) | 0.071<br>(0.023) | 0.001<br>(0.767) | 0.077<br>(0.014) | 0.003<br>(0.470) | 0.075<br>(0.017) |
| Withdrawal | **0.080\***<br>(*< .001*) | 0.060<br>(0.024) | 0.008<br>(0.126) | **0.132\***<br>(*< .001*) | 0.023<br>(0.014) | **0.118\***<br>(*< .001*) | 0.007<br>(0.316) | **0.133\***<br>(*< .001*) | 0.001<br>(0.768) | **0.139\***<br>(*< .001*) | 0.004<br>(0.472) | **0.136\***<br>(*< .001*) |
| Continued use | **0.095\***<br>(*< .001*) | 0.009<br>(0.773) | 0.013<br>(0.114) | 0.091<br>(0.005) | 0.035<br>(0.012) | 0.069<br>(0.021) | 0.008<br>(0.316) | **0.096\***<br>(0.003) | 0.000<br>(0.769) | **0.103\***<br>(0.002) | 0.004<br>(0.470) | **0.100\***<br>(0.002) |
| Hazardous use | **0.083\***<br>(*< .001*) | 0.012<br>(0.685) | 0.011<br>(0.116) | 0.085<br>(0.010) | 0.030<br>(0.012) | 0.065<br>(0.038) | 0.007<br>(0.316) | 0.088<br>(0.007) | 0.001<br>(0.768) | 0.095<br>(0.005) | 0.003<br>(0.471) | 0.092<br>(0.005) |
| Interpersonal problems | **0.094\***<br>(*< .001*) | 0.012<br>(0.686) | 0.013<br>(0.114) | **0.093\***<br>(0.004) | 0.033<br>(0.012) | 0.073<br>(0.015) | 0.008<br>(0.315) | **0.098\***<br>(0.002) | 0.000<br>(0.770) | **0.106\***<br>(0.001) | 0.003<br>(0.471) | **0.103\***<br>(0.002) |

Note. Sex, age, and 10 ancestry principal components were included as covariates. Observations were modeled as clustered within families. A Bonferroni correction for multiple testing was applied ( $p < .0045$ ). \* $p < .0045$

No significant mediation effects were observed for any personality psychopathology measure, including disconstraint (indirect effect *ps* > .012). This was notable given documented genetic correlations between consumption phenotypes and impulsive personality traits at the genome-wide level, which raised the possibility that the DPW PGS would show mediation through impulsivity even though it is nominally a narrower, consumption-specific score. As with the AUD PGS, several personality traits showed unique relations to AUD symptoms independent of the DPW PGS, indicating that these associations reflect processes independent of the genetic liability indexed by these PGS.

## Discussion

The present study examined whether genetic liabilities captured by AUD and DPW PGSs are expressed through alcohol consumption, personality psychopathology, or both, by testing mediation at the level of individual DSM-5 AUD symptoms rather than treating AUD as a single unitary outcome. Across nearly all individual symptoms, the relation between the AUD PGS and symptom expression was fully accounted for by heavier, habitual alcohol consumption (i.e., max drinks). Neuroticism also exhibited evidence of partially mediating the effect of the AUD PGS, while disconstraint (i.e., impulsivity) and aggressiveness did not, despite each of the latter showing unique associations with AUD. The relation between the DPW PGS and AUD symptoms, by contrast, operated exclusively through max drinks, with no mediating role for any personality dimension. Together, these results indicate that genetic liabilities captured by current AUD and DPW PGSs and their relations to AUD symptoms are largely accounted for by consumption levels, but the AUD PGS may also capture broader transdiagnostic influences, such as neuroticism, that are relevant to AUD.

Specifically, our findings suggest that the genetic liability underlying individual AUD symptoms may not be fully captured if they are treated as interchangeable indicators of a single underlying construct, as implicitly assumed by the DSM-5 (American Psychiatric Association, 2013). *Withdrawal* was the only symptom for which max drinks partially, rather than fully, mediated its relation with the AUD PGS. This could be due to withdrawal being among the more severe AUD criteria (Lane et al., 2016) and being associated with higher levels of genetic risk for AUD (Sarles et al., 2025). As such, *withdrawal* may reflect neuroadaptive changes that accumulate across repeated cycles of heavy drinking and abstinence, an allostatic process that is not reducible to drinking level alone (Koob & Volkow, 2016). Repeated withdrawal has been shown to produce lasting neuroadaptations in stress and emotional processing systems that persist into abstinence (Heilig et al., 2010). Furthermore, heavy drinkers who drink primarily for relief from negative states, including withdrawal, show greater AUD severity (Burnette et al., 2021). Therefore, the residual genetic association between the AUD PGS and *withdrawal* may reflect genetic influences related to susceptibility to this neuroadaptive process rather than to alcohol consumption per se.

Similarly, neuroticism also showed a distinct pattern of mediation when examining the relations of the AUD PGS to individual symptoms. It partially mediated the relation between the AUD PGS and nearly every symptom, but fully mediated its relation with *cut/stop* and *hazardous use*. One possible interpretation is that genetic liability for these two symptoms is more strongly expressed through affect-related processes than liability for other AUD symptoms. Watts et al. (2021) identified a multidimensional structure of AUD in which both *Cut/Stop* and *Hazardous Use* were represented within a broad Loss of Control dimension. Similarly, Kendler et al. (2012) identified a genetic factor characterized by loss of control and desire to quit drinking that was more strongly associated with major depressive disorder than other genetic factors underlying alcohol dependence criteria. These findings suggest that difficulties controlling alcohol use may involve genetic influences related to neuroticism. For example, *Cut/Stop* reflects unsuccessful attempts to reduce or control drinking, a process that affective models of addiction attribute substantially to negative reinforcement, in which the resumption of drinking is motivated by the desire to escape or avoid negative affect during attempted abstinence (Baker et al., 2004; Bresin et al., 2018).

Interestingly, the parallel mediation models provided additional context for these findings. The mediating effects of max drinks were largely unchanged when neuroticism was included as a second mediator, whereas the indirect effects through neuroticism were reduced by approximately half when max drinks was included in the same model. This attenuation suggests that the pathways through neuroticism and max drinks may not be entirely independent. One possibility is that neuroticism contributes to patterns of heavier alcohol consumption, for example through drinking to cope with negative affect (Kuntsche et al., 2006; Littlefield et al., 2011). In turn, some of its association with AUD symptoms thus operates through max drinks. Alternatively, neuroticism and max drinks may represent correlated but distinct phenotypes that account for overlapping variance in AUD symptoms. The present analyses were not designed to distinguish between these possibilities, and the cross-sectional mediation models cannot establish their causal ordering. Future research examining the temporal and etiological relations between neuroticism and max drinks may help clarify how these processes jointly contribute to individual AUD symptoms. Together, these findings support a more symptom-centered view of AUD genetic liability, in which the same aggregate genetic risk may be expressed through different etiological processes depending on the symptom under consideration.

In contrast to the AUD PGS, the DPW PGS showed a more consumption-specific pattern, operating exclusively through max drinks across all 11 AUD symptoms, with no mediation through neuroticism or any other personality psychopathology dimension. This pattern is in line with genomic evidence that alcohol consumption shows weaker genetic correlations with psychiatric and psychological traits than AUD or problematic alcohol use (Kranzler et al., 2019; Mallard et al., 2022; Walters et al., 2018). The absence of mediation through neuroticism for the DPW PGS therefore suggests that the neuroticism-related variance captured by the AUD PGS reflects genetic influences on the disorder that are not shared with level of consumption.

Finally, neither aggression nor impulsivity significantly mediated the relation between PGSs and AUD symptoms. For the AUD PGS, the absence of mediation through impulsivity is particularly noteworthy given previous PGS studies identifying mediating roles for sensation seeking and related impulsivity facets (Ksinan et al., 2019; Li et al., 2017; Su et al., 2021).

Aggression has also been implicated in the broader genetic liability shared between AUD and externalizing psychopathology (Barr et al., 2020; Waldman et al., 2020). Similarly, impulsivity did not mediate the relations between the DPW PGS and AUD symptoms, despite evidence of shared genetic influences between alcohol consumption and impulsivity (Sanchez-Roige et al., 2023; Stephenson et al., 2023). One possible explanation is that measurement differences may have contributed to these null findings. Impulsivity is a multifaceted construct (Dick et al., 2010), and genetic correlations across different self-report measures are generally moderate rather than complete (Sanchez-Roige et al., 2019; Stephenson et al., 2023). Thus, genetic associations identified using specific impulsivity measures, such as the scales of the UPPS-P (Mallard et al., 2026; Sanchez-Roige et al., 2019; Stephenson et al., 2023), may not translate directly to the broader behavioral disinhibition captured by the PSY-5 disconstraint scale.

Similarly, the PSY-5 aggressiveness scale may not capture the specific aspects of aggression most relevant to AUD genetic liability. Another possibility, particularly relevant to the AUD PGS, concerns the developmental differences in genetic influences on alcohol use. Kendler and his colleagues (2011) found that general externalizing genetic influences on alcohol behavior peak in mid-adolescence and then decline, whereas alcohol-specific genetic influences remain stable into adulthood. Because the present sample was ascertained for family history of AUD and assessed in adulthood (mean age = 50.04 years), it may disproportionately reflect this more persistent, alcohol-specific genetic risk relative to the developmentally earlier externalizing component.

Several limitations of this study should be considered. First, the sample was restricted to participants of European-like ancestry, which was necessary given the limited portability of PGSs across ancestry groups when discovery and target samples differ in ancestry composition (Martin et al., 2017; Otto et al., 2016). This limits the generalizability of the present findings to other populations. Second, impulsivity and aggression were each assessed using only a single variable. Therefore, future studies should examine whether alternative measures of impulsivity and aggression, including different facets of these constructs, show different patterns of mediation than those used in the present study. Third, the mediation models, while informative about statistical pathways, cannot establish causal ordering among the PGS, consumption, personality, and symptom variables. Fourth, the conservative Bonferroni correction, while appropriate for controlling family-wise error across 11 symptoms and multiple mediators, may have reduced power to detect smaller mediation effects, particularly for the personality psychopathology scales.

Despite these limitations, the present study has important implications for understanding the genetic liability captured by AUD and DPW PGSs. Specifically, the genetic liability indexed by an AUD PGS may not be equally relevant to all AUD symptoms. Although max drinks accounted for much of the association between both PGSs and AUD symptoms, *withdrawal* retained a direct association with the AUD PGS. This finding highlights the potential etiological heterogeneity of individual AUD symptoms and suggests that symptom-level analyses can reveal etiologically meaningful differences that may be obscured when AUD is treated as a single diagnostic phenotype. These findings also suggest the importance of conducting GWAS of individual AUD symptoms in addition to the AUD diagnosis and symptom counts alone. Such work is critical in advancing our understanding of the genetic architecture underlying AUD and helping to parse the pathways through which this genetic variation ultimately increases risk for AUD.

## Data Availability

Genotype data for the UCSF Family Study can be obtained through the Database for Genotypes and Phenotypes (dbGaP; Study Accession: phs001458.p1.v1). Phenotype data are available from the corresponding author upon request. PGC alcohol dependence GWAS summary statistics were obtained from the PGC website (https://www.med.unc.edu/pgc/). Million Veteran Program GWAS summary statistics were obtained through the Database for Genotypes and Phenotypes (dbGaP; Study Accession: phs001672). FinnGenR6 ICD-based AUD GWAS data were obtained from https://www.finngen.fi/en/access_results.

https://www.finngen.fi/en/access_results

## Acknowledgments

This study made use of summary statistics data from a number of sources. First, this study used GWAS summary statistics data from the sixth release of the FinnGen study. We thank the participants and investigators of the FinnGen study. Second, this research used summary data from the Psychiatric Genomics Consortium Substance Use Disorders (PGC-SUD) working group. PGC–SUD is supported by funds from NIDA and NIMH to MH109532 and, previously, had analyst support from NIAAA to U01AA008401 (COGA). PGC–SUD gratefully acknowledges its contributing studies and the participants in those studies, without whom this effort would not be possible. Third, this research used data from the Million Veteran Program and was supported by funding from the Department of Veterans Affairs Office of Research and Development, Million Veteran Program Grant nos. I01BX003341 and I01CX001849; and the VA Cooperative Studies Program study, no. 575B. This publication does not represent the views of the Department of Veterans Affairs or the United States Government.

