## Supplementary Table for "Examining the Etiological Specificity of Genetic Risk for Alcohol Use Disorder and Alcohol Consumption through Symptom-Level Mediation Analyses"

Supplementary Table 1. Associations between Mediators and Individual AUD Symptoms after Accounting for the AUD Polygenic Risk Score

| Dependent variable | Mediators | | | | | |
|---|---|---|---|---|---|---|
| | Maximum habitual drinks | Aggressiveness | Disconstraint | Neuroticism | Introversion | Psychoticism |
| | Effect from mediator to symptom *(p)* | Effect from mediator to symptom *(p)* | Effect from mediator to symptom *(p)* | Effect from mediator to symptom *(p)* | Effect from mediator to symptom *(p)* | Effect from mediator to symptom *(p)* |
| Cut/Stop | **0.080*** *(< .001)* | **0.021*** *(< .001)* | **0.027*** *(< .001)* | **0.022*** *(< .001)* | 0.005 *(0.040)* | **0.015*** *(< .001)* |
| Larger /longer | **0.086*** *(< .001)* | **0.026*** *(< .001)* | **0.040*** *(< .001)* | **0.025*** *(< .001)* | 0.006 *(0.038)* | **0.015*** *(< .001)* |
| Craving | **0.069*** *(< .001)* | **0.017*** *(< .001)* | **0.023*** *(< .001)* | **0.025*** *(< .001)* | 0.005 *(0.073)* | **0.016*** *(< .001)* |
| Time spent | **0.074*** *(< .001)* | **0.028*** *(< .001)* | **0.032*** *(< .001)* | **0.024*** *(< .001)* | 0.008 *(0.005)* | **0.016*** *(< .001)* |
| Tolerance | **0.092*** *(< .001)* | **0.030*** *(< .001)* | **0.041*** *(< .001)* | **0.029*** *(< .001)* | 0.007 *(0.008)* | **0.019*** *(< .001)* |
| Activities given up | **0.067*** *(< .001)* | **0.017*** *(< .001)* | **0.028*** *(< .001)* | **0.025*** *(< .001)* | **0.011*** *(< .001)* | **0.016*** *(< .001)* |
| Role failure | **0.068*** *(< .001)* | **0.018*** *(< .001)* | **0.030*** *(< .001)* | **0.025*** *(< .001)* | **0.009*** *(0.001)* | **0.015*** *(< .001)* |
| Withdrawal | **0.077*** *(< .001)* | **0.019*** *(< .001)* | **0.032*** *(< .001)* | **0.024*** *(< .001)* | **0.008*** *(0.003)* | **0.017*** *(< .001)* |
| Continued use | **0.092*** *(< .001)* | **0.032*** *(< .001)* | **0.048*** *(< .001)* | **0.027*** *(< .001)* | 0.005 *(0.091)* | **0.017*** *(< .001)* |
| Hazardous use | **0.080*** *(< .001)* | **0.025*** *(< .001)* | **0.042*** *(< .001)* | **0.024*** *(< .001)* | 0.005 *(0.050)* | **0.015*** *(< .001)* |
| Interpersonal problems | **0.090*** *(< .001)* | **0.031*** *(< .001)* | **0.046*** *(< .001)* | **0.027*** *(< .001)* | 0.003 *(0.188)* | **0.016*** *(< .001)* |

*Note.* Sex, age, and 10 ancestry principal components were included as covariates. Observations were modeled as clustered within families. A Bonferroni correction for multiple testing was applied ($p < .0045$). *p < .0045

Supplementary Table 2. Results of Parallel Mediation Analyses for the AUD Polygenic Risk Score

| Dependent variable | Mediators | | |
|---|---|---|---|
| | | Maximum habitual drinks | Neuroticism |
| | Direct effect (*p*) | Indirect effect (*p*) | Indirect effect (*p*) |
| Cut/Stop | 0.026 *(0.389)* | **0.069*** *(< .001)* | 0.010 *(0.013)* |
| Larger /longer | 0.043 *(0.169)* | **0.074*** *(< .001)* | 0.012 *(0.009)* |
| Craving | 0.069 *(0.016)* | **0.059*** *(< .001)* | **0.015*** *(0.004)* |
| Time spent | 0.047 *(0.097)* | **0.064*** *(< .001)* | 0.013 *(0.006)* |
| Tolerance | 0.044 *(0.159)* | **0.079*** *(< .001)* | **0.016*** *(0.004)* |
| Activities given up | 0.060 *(0.045)* | **0.057*** *(< .001)* | **0.016*** *(0.003)* |
| Role failure | 0.057 *(0.051)* | **0.057*** *(< .001)* | 0.015 *(0.005)* |
| Withdrawal | **0.081*** *(0.003)* | **0.066*** *(< .001)* | 0.013 *(0.006)* |
| Continued use | 0.047 *(0.139)* | **0.079*** *(< .001)* | 0.014 *(0.007)* |
| Hazardous use | 0.027 *(0.417)* | **0.069*** *(< .001)* | 0.012 *(0.011)* |
| Interpersonal problems | 0.046 *(0.142)* | **0.078*** *(< .001)* | 0.014 *(0.007)* |

*Note.* Sex, age, and 10 ancestry principal components were included as covariates. Observations were modeled as clustered within families. A Bonferroni correction for multiple testing was applied ($p < .0045$).
*p < .0045

Supplementary Table 3. Associations between Mediators and Individual AUD Symptoms after Accounting for the DPW Polygenic Risk Score

| Dependent variable | Mediators | | | | | |
|---|---|---|---|---|---|---|
| | Maximum habitual drinks | Aggressiveness | Disconstraint | Neuroticism | Introversion | Psychoticism |
| | Effect from mediator to symptom *(p)* | Effect from mediator to symptom *(p)* | Effect from mediator to symptom *(p)* | Effect from mediator to symptom *(p)* | Effect from mediator to symptom *(p)* | Effect from mediator to symptom *(p)* |
| Cut/Stop | **0.081*** | **0.021*** | **0.027*** | **0.023*** | 0.005 | **0.014*** |
| | *(< .001)* | *(< .001)* | *(< .001)* | *(< .001)* | *(0.040)* | *(< .001)* |
| Larger /longer | **0.087*** | **0.025*** | **0.040*** | **0.026*** | 0.006 | **0.015*** |
| | *(< .001)* | *(< .001)* | *(< .001)* | *(< .001)* | *(0.036)* | *(< .001)* |
| Craving | **0.070*** | **0.016*** | **0.023*** | **0.025*** | 0.005 | **0.016*** |
| | *(< .001)* | *(< .001)* | *(< .001)* | *(< .001)* | *(0.073)* | *(< .001)* |
| Time spent | **0.075*** | **0.027*** | **0.032*** | **0.024*** | **0.008*** | **0.016*** |
| | *(< .001)* | *(< .001)* | *(< .001)* | *(< .001)* | *(0.004)* | *(< .001)* |
| Tolerance | **0.092*** | **0.030*** | **0.041*** | **0.030*** | 0.007 | **0.019*** |
| | *(< .001)* | *(< .001)* | *(< .001)* | *(< .001)* | *(0.007)* | *(< .001)* |
| Activities given up | **0.068*** | **0.017*** | **0.028*** | **0.026*** | **0.010*** | **0.016*** |
| | *(< .001)* | *(< .001)* | *(< .001)* | *(< .001)* | *(< .001)* | *(< .001)* |
| Role failure | **0.068*** | **0.017*** | **0.029*** | **0.025*** | **0.009*** | **0.015*** |
| | *(< .001)* | *(< .001)* | *(< .001)* | *(< .001)* | *(< .001)* | *(< .001)* |
| Withdrawal | **0.077*** | **0.019*** | **0.031*** | **0.025*** | **0.008*** | **0.018*** |
| | *(< .001)* | *(< .001)* | *(< .001)* | *(< .001)* | *(0.003)* | *(< .001)* |
| Continued use | **0.092*** | **0.031*** | **0.048*** | **0.028*** | 0.005 | **0.017*** |
| | *(< .001)* | *(< .001)* | *(< .001)* | *(< .001)* | *(0.088)* | *(< .001)* |
| Hazardous use | **0.080*** | **0.025*** | **0.042*** | **0.024*** | 0.006 | **0.015*** |
| | *(< .001)* | *(< .001)* | *(< .001)* | *(< .001)* | *(0.036)* | *(< .001)* |
| Interpersonal problems | **0.091*** | **0.030*** | **0.046*** | **0.028*** | 0.004 | **0.016*** |
| | *(< .001)* | *(< .001)* | *(< .001)* | *(< .001)* | *(0.183)* | *(< .001)* |

*Note.* Sex, age, and 10 ancestry principal components were included as covariates. Observations were modeled as clustered within families. A Bonferroni correction for multiple testing was applied ($p < .0045$). *p < .0045
